# Clinical Decisions Without EEG: High Anti-Seizure Medication Use Before EEG in Infants and Its Implications

**DOI:** 10.1101/2025.05.09.25326226

**Authors:** Nathalia Beller, Madeline Fields, Chad H Hogan, Sushma Krishna, Benjamin S Glicksberg, Courtney E Juliano, Felix Richter

**Author notes:** **Corresponding Author** Felix Richter, MD PhD.

## Abstract

Neonatal seizures are challenging to diagnose due to subtle clinical presentations and require video-EEG for confirmatory diagnosis. However, video-EEG is not always available, so clinicians must sometimes decide to treat neonatal seizures prior to diagnostic confirmation. The extent of this gap in clinical care is not previously described. This retrospective study examined anti-seizure medication (ASM) use in 115 infants who underwent video-EEG at Mount Sinai. Of 46 infants treated with ASMs, 59% received loading doses before EEG. Among these, 89% showed epileptiform activity and 52% (14/27) had seizures on EEG. Mortality among treated infants was high (30%). These findings highlight the significant reliance on clinical judgment without EEG when treating neonatal seizures. Our data support future work to develop scalable, accessible tools to improve timely and accurate treatment of neonatal seizures.

---

Neonatal seizures are among the most common neurologic emergencies in newborns. These events can be difficult to detect due to their often subtle and varied clinical presentations, which can be mistaken for normal neonatal movements or other benign conditions such as jitteriness or sleep myoclonus.^1^ Accurate seizure identification is crucial, as untreated seizures may lead to brain injury, while unnecessary treatment exposes infants to the risks of anti-seizure medications (ASMs), including drug toxicity and adverse developmental effects.^2^

Video electroencephalography (EEG) is the gold standard for neonatal seizure detection, yet its cost and need for specialized expertise limit its widespread use.^3^ Understanding the gaps in care due to restricted video-EEG availability is critical to improving access or developing new technology to fill these gaps in neonatal care. While prior studies have examined treatment delays after EEG initiation,^4,5^ none have assessed ASM use before EEG confirmation. Our study provides real-world data on ASM administration rates based on clinical suspicion alone (i.e., without EEG), associated underlying neuropathology, the context of initial care where seizures were first suspected (e.g., an outside hospital without video-EEG prior to transfer), and the ultimate outcome of seizures and epileptiform activity. The results highlight gaps in neonatal seizure management and the need for more accessible diagnostic tools.

Clinical data were retrospectively collected between Feb 2021 and Dec 2022 for 115 infants under one year of age who underwent video-EEG monitoring at Mount Sinai Kravis Children’s Hospital. The Institutional Review Board at the Icahn School of Medicine at Mount Sinai approved this study. A total of 45 (39%) of infants received a loading dose of at least one ASM (**Supplemental Table 1**). We observed that 59% (27/46) of infants received ASM loading doses before undergoing video-EEG, while 41% (19/46) received ASMs only after starting video-EEG monitoring. Of the infants who received ASMs prior to EEG, 89% (24/27) had epileptiform activity including 52% (14/27) with seizures subsequently observed on EEG (**Figure 1**). This suggests an upper bound on the false positive rate of seizure detection of 48% based on clinical suspicion alone without EEG, although this false positive rate could be lower if some infants with epileptiform activity on EEG had seizures prior to treatment.

**Figure 1.**
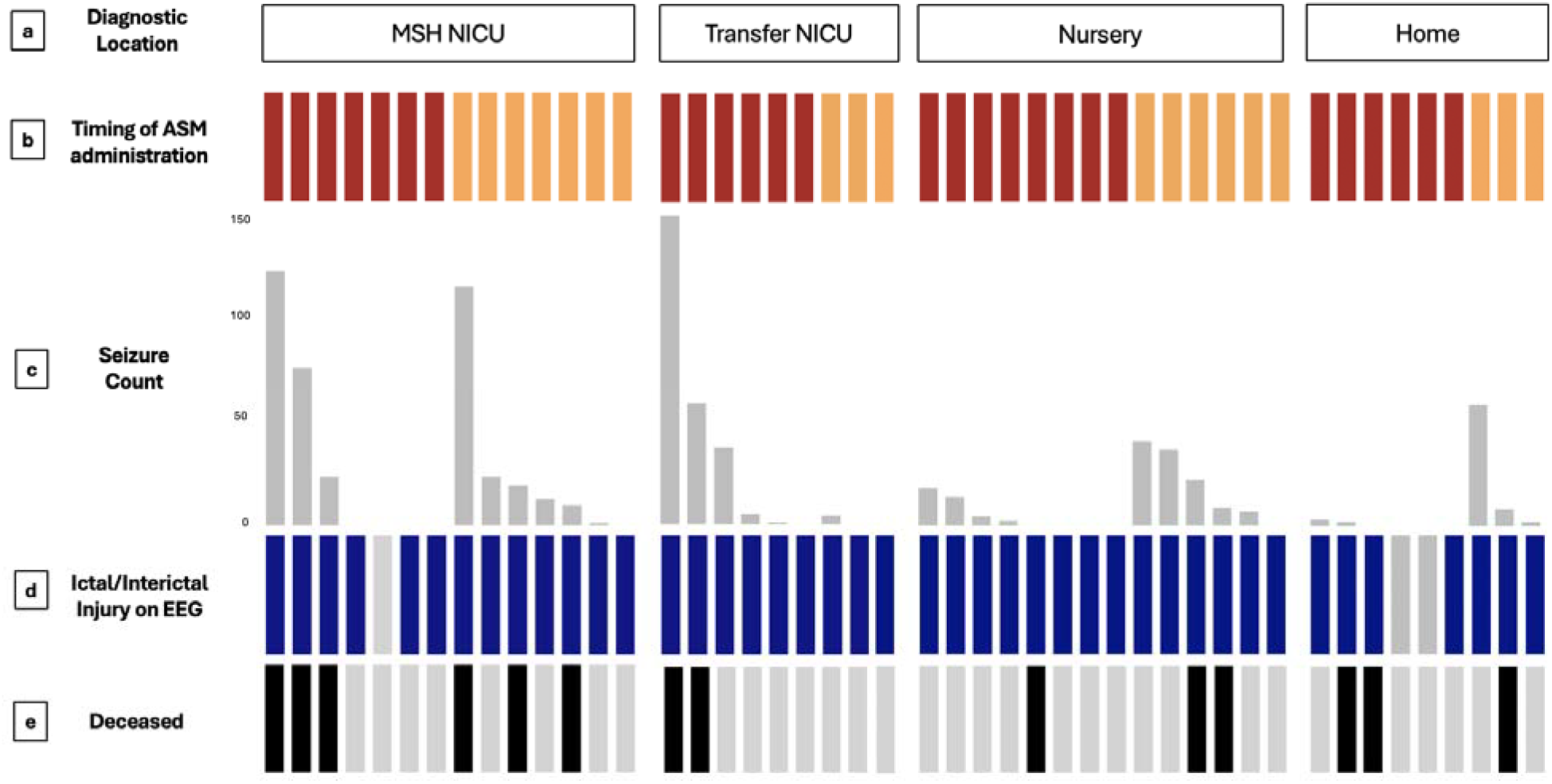
Seizure detection among the 46 infants who received ASM loading doses and video-EEG. a) Initial clinical context in which a concerning neurological finding or seizure risk factor was first observed. Notably, 17.8% occurred at a NICU without video-EEG (‘Transfer NICU’), 31.1% occurred in the well-baby nursery, and 20.0% were observed at home. **b)** Timing of anti-seizure medication (ASM) administration relative to when video-EEG was initiated. Red represents infants who received ASM prior to video-EEG hookup. **c)** Number of seizures detected on video-EEG per infant. **d)** Presence of epileptiform activity on video-EEG (dark blue) indicating ictal or interictal injury, which could be a reflection of prior seizure activity or proclivity to future seizures. **e)** Mortality, with black depicting infants who ultimately passed away. Acronyms: MSH = Mount Sinai Hospital; NICU = Neonatal Intensive Care Unit; ASM = anti-seizure medication; EEG = electroencephalogram

Phenobarbital, either by itself or in combination with other medications, was used most frequently (52%; 24/46), followed by Levetiracetam (26%; 12/46). The distribution of neuropathology was diverse among this population (**Supplemental Table 1**). IVH appeared more common among infants who received ASMs prior to EEG initiation (18.5% vs. 0%, Fisher’s exact test P=0.067), while HIE showed a trend toward greater prevalence in those who received ASMs only after EEG was initiated (OR=3.4, P=0.15); however, neither association reached statistical significance. Mortality was high (30%; 14/46) among all infants who received ASM loading doses, highlighting their acuity, and was similar in both those who were treated before versus after starting video-EEG (OR=0.91, P=1). Clinical suspicion for seizures that prompted treatment before EEG occurred most frequently in the NICU (48.1%; 13/27), followed by the nursery (29.6%; 8/27) and the home (22.2%; 6/27). All infants underwent video-EEG in the NICU, except those in the ‘home’ group who underwent video-EEG in the pediatric wards or ICU. Many of these infants (37%, 10/27) were transferred to Mount Sinai from outside NICUs that did not have access to video-EEG. This finding highlights the frequency with which neurologic emergencies are treated without EEG support in the United States.^4^

Seizure management remains a critical challenge in neonatology, due to the difficulty in distinguishing seizures from non-epileptic movements, and the reliance on clinical suspicion in settings where video-EEG is not immediately available.^2,3^ While other studies have investigated seizure treatment delays after starting EEG,^4,5^ ours is among the first real-world datasets to directly measure the high rate of decisions to administer ASMs that occurs *without* EEG. The frequent administration of ASMs before EEG confirmation emphasizes the need for scalable diagnostic tools in the diversity of healthcare settings in which infants can present with seizures. Recent advances in AI present new opportunities to address these limitations. AI decision support is already used with video-EEG to facilitate analysis,^6,7^ however portable EEG devices for automated seizure detection in infants have not yet seen significant uptake or regulatory approval. In addition, computer vision approaches could augment EEG devices either with artifact removal or semiology classification,^8^ especially with focal seizures or rarer genetic epilepsy subtypes. Further research is needed to refine seizure identification methods to provide more timely interventions, reduce strain on neurologic emergency transport systems, and ultimately improve outcomes for this devastating neurologic emergency in neonates and infants.

## Supporting information

Supplemental Material: ASM use before EEG

## Data Availability

All data produced in the present study are available upon reasonable request to the authors.

