## Supplemental Material: ASM use before EEG for "Clinical Decisions Without EEG: High Anti-Seizure Medication Use Before EEG in Infants and Its Implications"

**Running title**
Anti-Seizure Medications before EEG in Infants

**Authors**
Nathalia Beller[1], Madeline Fields[2], Chad H Hogan[1], Sushma Krishna[3], Benjamin S Glicksberg[4,5,6,7], Courtney E Juliano[3], Felix Richter[3,4,5,7]

**Affiliations**
[1]Department of Genetics, Icahn School of Medicine at Mount Sinai, New York, NY. [2]Department of Neurology, Icahn School of Medicine at Mount Sinai, New York, NY. [3]Division of Neonatology, Department of Pediatrics, Icahn School of Medicine at Mount Sinai. [4]Mindich Child Health and Development Institute, Icahn School of Medicine at Mount Sinai, New York, NY. [5]Department of Artificial Intelligence, Icahn School of Medicine at Mount Sinai, New York, NY. [6]Department of Pediatrics, Icahn School of Medicine at Mount Sinai, New York, NY. [7]Center for AI in Children's Health, Icahn School of Medicine at Mount Sinai, New York, NY.

**Corresponding Author**
Felix Richter, MD PhD


**Manuscript Number** MEDRXIV/2025/326226

**<u>Supplemental Table 1</u>:** Demographic and clinical characteristics of all study infants.

| Descriptive factor | | Before vEEG N = 27.0 | After vEEG N = 19.0 | Infants with ASM load N = 46 | Infants without ASM load N = 69 | All Infants N = 115 |
|---|---|---|---|---|---|---|
| Gestational Age at Birth, median (range) | | 36.9w* (24.6w - 41.1w) | 37.8w (30.4w - 40.1w) | 37.3 (24.1w - 41.1w) | 36.3 (23.0w - 41.0w) | 36.7 (23.0w - 41.1w) |
| | < 1 | 20 (74.1) | 14 (73.7) | 34 (73.4) | 43 (62.3) | 77 (67.0) |
| | [1 - 3) | 5 (18.5) | 4 (21.0) | 9 (19.6) | 13 (18.8) | 22 (19.1) |
| Age in months of first ASMs | [3 - 6) | 1 (3.7) | 0 (00.0) | 1 (2.2) | 8 (11.6) | 9 (7.8) |
| administration, *N (%)* | ≥ 6 | 1 (3.7) | 1 (5.2) | 2 (4.3) | 5 (7.2) | 7 (6.1) |
| | Female | 12 (44.4) | 7 (36.8) | 19 (41.3) | 34 (49.3) | 53 (46.1) |
| Sex, *N (%)* | Male | 15 (55.6) | 12 (63.2) | 27 (58.7) | 35 (50.7) | 62 (53.9) |
| | American Indian or Alaskan | 2 (7.4) | 2 (10.5) | 4 (8.7) | 1 (1.4) | 5 (4.3) |
| | Asian | 2 (7.4) | 1 (5.2) | 3 (6.52) | 7 (10.1) | 10 (8.7) |
| | Black or African-American | 5 (18.5) | 4 (21.5) | 9 (19.6) | 18 (26.1) | 27 (23.5) |
| | Native Hawaiian or Pacific Islander | 0 (00.0) | 0 (00.0) | 0 (00.0) | 1 (1.4) | 1 (0.9) |
| | White | 6 (22.2) | 8 (42.1) | 14 (30.4) | 17 (24.6) | 31 (27.0) |
| | Other** | 8 (29.6) | 3 (15.8) | 11 (23.9) | 22 (31.9) | 33 (29.0) |
| Race, *N (%)* | Unknown | 4 (14.8) | 1 (5.2) | 5 (10.9) | 3 (4.3) | 8 (7.0) |
| | Hispanic/Latino | 9 (33.3) | 4 (21.5) | 13 (28.3) | 21 (30.4) | 34 (29.6) |
| | Non-Hispanic | 11 (40.7) | 12 (63.2) | 23 (50.0) | 35 (50.7) | 58 (50.4) |
| Ethnicity, *N (%)* | Unknown | 7 (25.9) | 3 (15.8) | 10 (21.7) | 13 (18.8) | 23 (20.0) |
| | Hypoxic ischemic encephalopathy | 4 (14.8) | 7 (36.8) | 11 (23.9) | 14 (20.3) | 25 (21.7) |
| | Genetic or idiopathic epilepsy | 5 (18.5) | 5 (26.3) | 10 (21.7) | 4 (5.8) | 14 (12.2) |
| | Intraventricular hemorrhage | 5 (18.5) | 0 (00.0) | 5 (10.9) | 2 (2.9) | 7 (6.1) |
| | Stroke | 3 (11.1) | 2 (10.5) | 5 (10.9) | 0 (00.0) | 5 (4.3) |
| | Structural brain malformation | 2 (7.4) | 2 (10.5) | 4 (8.7) | 1 (1.4) | 5 (4.3) |
| | Other | 5 (18.5) | 3 (15.8) | 8 (17.4) | 7 (10.1) | 15 (13.0) |
| Neurologic pathology, *N (%)* | None | 3 (11.1) | 0 (00.0) | 3 (6.5) | 41 (59.4) | 44 (38.3) |

*w, weeks.

**Other, option if the caregiver did not self-identify with a pre-specified racial category.
